# Shared genetic architecture of cortical morphology and psychiatric disorders: insights from a cross-trait analyses across 180 cortical regions

**DOI:** 10.64898/2026.04.10.26349224

**Authors:** Yingzhe Zhang, Tian Ge, Travis T. Mallard, Karmel W Choi, Anxiety Disorders Working Group of the Psychiatric Genomics Consortium, Henning Tiemeier, Sander Lamballais

## Abstract

The shared genetic liability between cortical morphology and psychiatric disorders remains unclear. We aimed to identify whether the genetic loci shared between cortical morphology and six psychiatric disorders show regional or global effects. We identified substantial pairwise genetic overlaps of cortical surface area or thickness with psychiatric disorders; however, these loci lacked a uniform direction (∼50% concordance). Cross-trait analyses revealed distinct architectures: internalizing disorders and schizophrenia/bipolar disorder shared more genetic loci with localized effects, whereas neurodevelopmental disorders shared fewer loci but more with widespread effects. We identified 17 genomic loci shared across all disorders, most of which had opposing directional effects across cortical regions. Only one locus (rs2431112) had region-specific and unidirectional effects (reduced primary visual and posterior cingulate surface area). This directional heterogeneity within and across pleiotropic loci reveals complex shared genetic architectures and likely limits the genetic predictive performance of brain morphology for psychiatric disorders.

## Introduction

Psychiatric disorders exhibit substantial transdiagnostic phenotypic overlap, with shared clinical features and comorbidities observed across diagnostic categories.^1–3^ Comorbidity is highly prevalent, with about half of psychiatric patients presenting with multiple diagnoses.^4^ Accumulating evidence demonstrates significant genetic correlations between psychiatric disorders, with genome-wide association studies (GWASs) revealing extensive pleiotropy. On average, cross-disorder GWAS analyses have reported a genetic correlation of approximately 0.4 among psychiatric conditions,^5^ with particularly high correlations observed between schizophrenia and bipolar disorder (rg = 0.68), and between anxiety and major depressive disorders (rg = 0.91).^6–8^ These genetic overlaps provide a compelling biological explanation for the observed transdiagnostic phenomena, suggesting that shared genetic architectures contribute to the phenotypic overlap between psychiatric disorders.^9–14^

Neuroimaging studies have identified numerous brain structural correlates of psychiatric disorders, though these findings often lack disease specificity and consistency across studies.^15^ Large-scale analyses from consortiums like ENIGMA have revealed that major depressive disorder, bipolar disorder, schizophrenia, and obsessive-compulsive disorder share correlated patterns of cortical abnormalities, particularly in the prefrontal cortex and anterior cingulate.^16,17^ However, the associated neuroanatomical alterations are typically widespread, nuanced, and not specific for any given diagnosis.^15,18,19^ The observation that the relationships tend to be complex and transdiagnostic in nature raises the question whether common underlying biological mechanisms across psychiatric disorders drive the shared neural signatures. Shared genetic factors might simultaneously affect multiple brain regions, producing overlapping patterns of structural alterations across different psychiatric conditions.^20–22^

Genetic research offers crucial advantages in uncovering the biological mechanisms underlying the associations between brain structures and psychiatric disorders, as genetic variants are not susceptible to reverse causation or environmental confounding. Emerging work has begun to probe the shared genetic architecture between brain structures and single psychiatric disorders.^23,24^ However, pairwise cross-trait analyses are limited in that they cannot simultaneously model relationships across multiple psychiatric disorders and multiple brain regions. Consequently, it remains unclear whether genetic loci affecting several psychiatric disorders exert region-specific influences or are systematically associated with widespread brain structures. Clarifying these patterns is essential for disentangling the biological pathways linking brain structure and psychopathology, and for advancing precision psychiatry approaches.

In this study, we investigated the shared genomic loci affecting both cortical structure and six psychiatric disorders by leveraging large-scale GWAS summary statistics. Our aim was to systematically identify shared common genetic loci linking either regional or global cortical signal by jointly modeling individual psychiatric disorders, psychiatric dimensions and general psychopathology with 180 brain region morphology.

## Results

### Study overview

We leveraged large-scale GWAS data on total and regional (180 bilaterally averaged regions) cortical surface area and thickness, along with six psychiatric disorders: attention-deficit/hyperactivity disorder (ADHD), autism spectrum disorder (ASD), anxiety, bipolar disorder (BP), major depressive disorder (MDD), and schizophrenia (SCZ). First, we quantified the extent and directional patterns of overlapping causal variants between overall brain structure and each psychiatric disorder using bivariate causal mixture models (MiXeR). Next, we identified genetic variants influencing both brain morphology and psychiatric disorders with SumRank, a cross-trait association method that integrates GWAS p-values across multiple traits to determine, for each variant, the subset of traits to which it is most strongly associated.^25^ SumRank can robustly control the false positive rate while being well-powered to detect cross-trait associations across hundreds of traits. We applied SumRank to the summary statistics for the cortical thickness and surface area of 180 brain regions and psychiatric disorders in three stages, for each individual psychiatric disorder, psychiatric disorder dimensions defined by the cross-disorder GWAS study (neurodevelopmental disorders, internalizing disorders and schizophrenia/bipolar disorders)^5^, and all six psychiatric disorders jointly. Finally, we performed functional annotation of these shared variants and conducted gene set analyses to gain insight into the underlying biological mechanisms linking single and multiple disorders to cortical structures.

### Quantification of genetic overlap between overall brain morphology and each psychiatric disorder

Using univariate MiXeR, we estimated heritability, polygenicity, and discoverability for overall surface area and cortical thickness as well as for each psychiatric disorder. All traits showed good model fit, indicated by positive Akaike Information Criterion values. Overall surface area had an observed SNP heritability of 26.7% with 1,651 influencing variants and a discoverability of 2.9×10⁻⁴. Such low discoverability indicated that causal SNPs have very small effects on overall surface are and spread thinly across genome. Overall cortical thickness showed an observed SNP heritability of 32.8%, influenced by 1,742 variants with a discoverability of 2.5×10⁻⁴. For the psychiatric disorders, observed SNP heritability estimates ranged from 3% for anxiety and depression to 18.5% for ASD and 37.1% for schizophrenia (Supplementary Data 1). These traits were influenced by between 6,079 and 11,190 variants, with discoverability estimates spanning from 4.21×10⁻⁶ for depression to 5.94×10⁻⁵ for schizophrenia.

Bivariate MiXeR showed that overall brain morphology shared substantial causal variants with individual psychiatric disorders, as illustrated in Figure 1. Of the variants related to total surface area, we found the overlap with variants related to psychiatric disorders ranging from 38.9% for ASD to 94.4% with schizophrenia. For cortical thickness, overlap ranged from 50.0% for ADHD to 94.1% for anxiety and schizophrenia. Conversely, the variants related to psychiatric disorders shared a smaller proportion with cortical structures (for cortical thickness this ranged from 8.9% for depression to 19.7% for ASD). Full results are provided in Supplementary Data 2.

**Figure 1.**
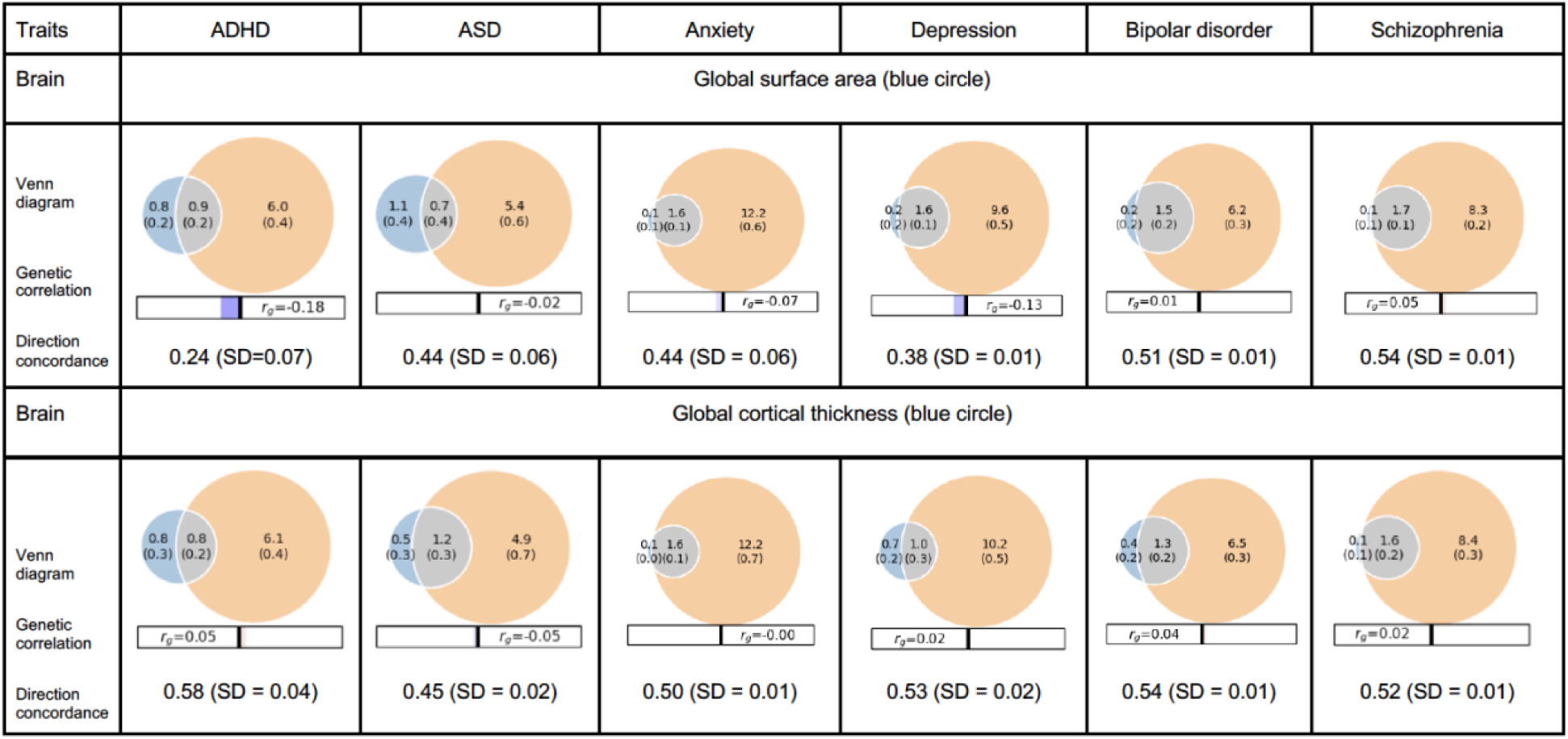
Overlap of causal genetic variants for global brain structures and specific psychiatric disorders assessed with bivariate gaussian mixture modeling. The orange circle on the right represents causal variants for the specific psychiatric disorder, the blue circle on the left represents the causal variants for global surface area or global cortical thickness, and the grey overlap area represents the shared causal variants. The numbers indicate the estimated quantity of causal variants (in thousands) per component, explaining 90% of SNP heritability in each phenotype, followed by the standard error.

Next, we calculated the directional concordance of the shared genetic variants. This corresponds to the sign of the respective genetic correlations for each trait pair. Note, the larger the difference to 50%, the stronger the pattern of directionality. The overall brain morphology and psychiatric disorders mostly exhibited divergent directions of effect. ASD, bipolar disorder and schizophrenia each showed around 50% (i.e. random) concordance in direction with the overlapping causal variants related to total surface area. Variants signaling a higher risk for ADHD and depression demonstrated lower concordance with variants related to more total surface area (24% and 38%, respectively). All six psychiatric disorders showed a concordance with total cortical thickness consistent with chance expectation (around 50%, i.e. no clear directional pattern despite substantial genetic overlaps). For comparison, we performed bivariate MiXeR analyses across psychiatric disorders only. These analyses revealed substantial shared genetic architectures (26–100% causal variants overlap) accompanied by significant positive genetic correlations (r = 0.23–0.89). In contrast to the analyses for brain morphology and psychiatric disorders, the shared causal variants between psychiatric disorders showed high directional concordance, with a mean of 79%, indicating largely consistent directions (Supplementary Data 3). This contrast indicates that shared causal variants are directionally aligned among psychiatric disorders, but not between brain morphology and psychiatric disorders.

### Cross-traits shared genetics between specific brain regions and psychiatric disorders

For each psychiatric disorder, we used SumRank to identify genetic variants associated both with the disorder and with the surface area or thickness of at least one cortical region, at a genome-wide significant threshold of 5 × 10^−8^. For cortical surface area, we identified 194 shared independent SNPs shared with ASD, 453 with ADHD, 539 with anxiety, 678 with bipolar disorder, 1,153 with MDD, and 1,364 with schizophrenia (details in Supplementary Data 4-9). For cortical thickness, the number of overlapping SNPs were 352 for ASD, 704 for ADHD, 769 for anxiety, 1,022 for bipolar disorder, 1,694 for MDD, and 1,978 for schizophrenia (details in Supplementary Data 10-15).

The distribution of the number of brain regions associated with the overlapping independent SNPs varied widely for each disorder. We plotted the density of all significant independent SNPs per disorder as a function of the number of related brain regions (Figure 2A). Absolute SNP count distributions are provided in Supplementary Figure 1. For surface area and cortical thickness, overlapping variants affected 1–41 regions, with a median of 2 regions per each psychiatric disorder except for ASD. For ASD, each locus shared with surface area was linked to a median of 3 regions (IQR 1–11), and each locus shared with cortical thickness was linked to a median of 4 regions (IQR 2–11). (Figure 2B).

**Figure 2.**
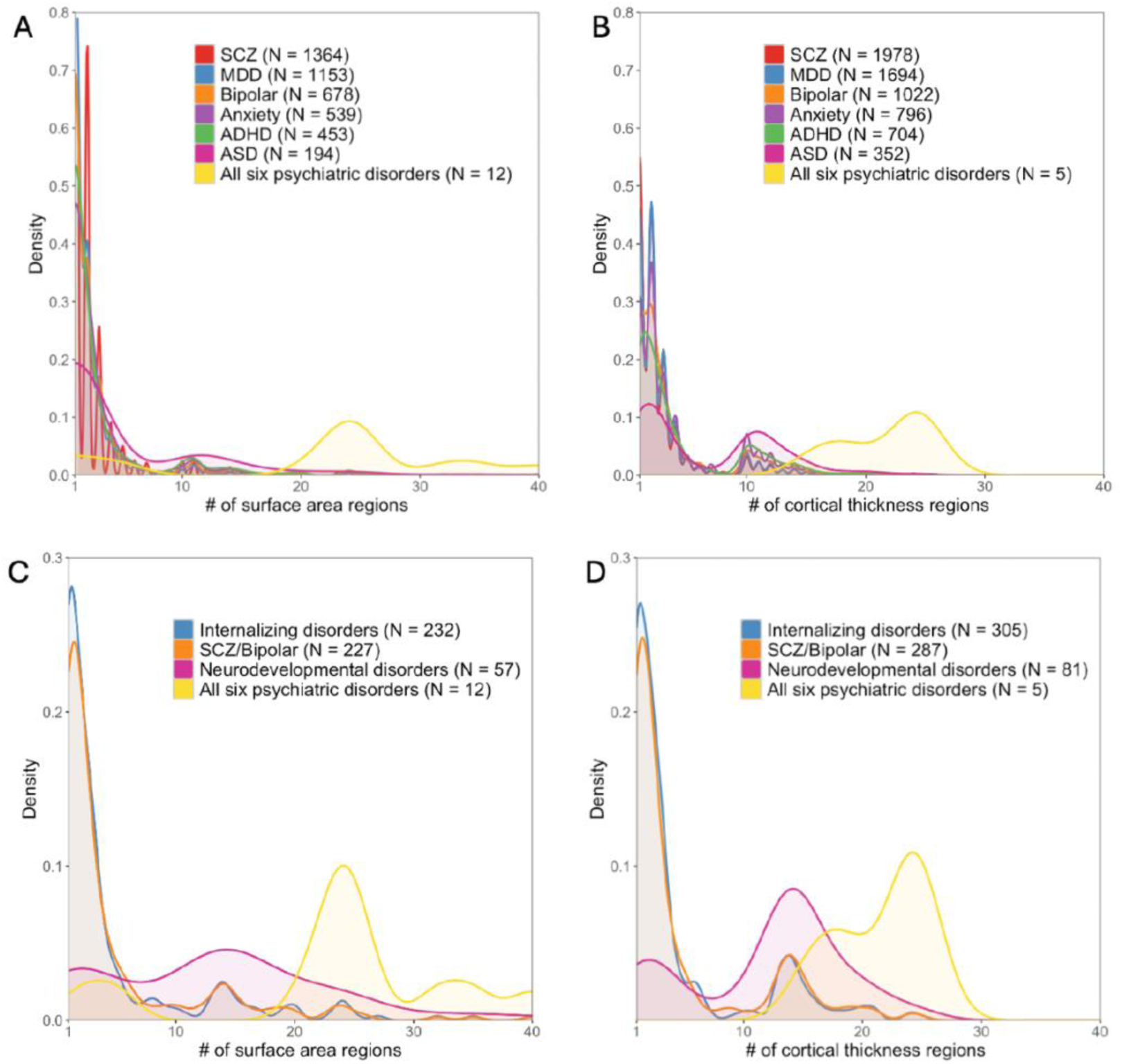
Distribution of genetic variants shared by a certain number of brain regions (cortical thickness and area) with psychiatric traits: Different patterns for specific psychiatric disorders, psychiatric disorder groups, and general psychopathology. N in the figures showed the number of significant independent SNPs identified between one or more of 180 brain regions and each psychiatric disorder, psychiatric dimensions and general psychopathology (all six psychiatric disorders).

Next, we restricted analyses to traits that included at least one brain region and the psychiatric disorders within the psychiatric disorder dimensions defined by the cross-disorder GWAS study.^5^ For neurodevelopmental disorders, we identified 57 independent SNPs associated with a median of 14 surface area regions (IQR = [3,19]). In contrast, for internalizing disorders, 232 SNPs were associated with a median of 2 regions (IQR = [1,4]), and for schizophrenia/bipolar disorders, 227 SNPs mapped to a median of 2 regions (IQR = [1,5]). Details were included in Supplementary Data 16-18). Again, we found a very similar pattern for cortical thickness (Figure 2C,2D, Supplementary Data 19-21).

### Cross-traits shared genetics between brain regions and general psychopathology

When examining variants associated with cortical surface area of any of the 180 regions and all six psychiatric disorders (i.e., “general psychopathology”), 12 independent genomic loci were identified. These loci were linked to the surface area of 2 to 40 brain regions (median = 24, IQR [22.5,27.5]). An interesting pattern emerged: most independent SNPs were associated with multiple brain regions but exhibited directionally heterogeneous effects across regions. Only three SNPs had consistent effect directions for surface area (Table 1). Of these, variant rs2431112 (near *NIHCOLE* and *RNU6-334P*; p = 6.38×10⁻³²) showed consistent negative associations with surface area in the primary visual cortex and dorsal area 23a+b, and was associated with reduced risk across all six psychiatric disorder. Variant rs62057151, previously associated with worrying,^26^ was consistently positively associated with surface area across 21 regions. Similarly, variant rs8039305 (intron of *FURIN*; p = 2.52×10⁻⁸) was positively associated with ADHD, anxiety, bipolar disorder, MDD, and schizophrenia in their original GWASs, and demonstrated positive associations with several regions, including area OP4/PV, frontal opercular area 2, precuneus visual area, area TF in ventral temporal lobe, and lateral intraparietal ventral.

**Table 1.**
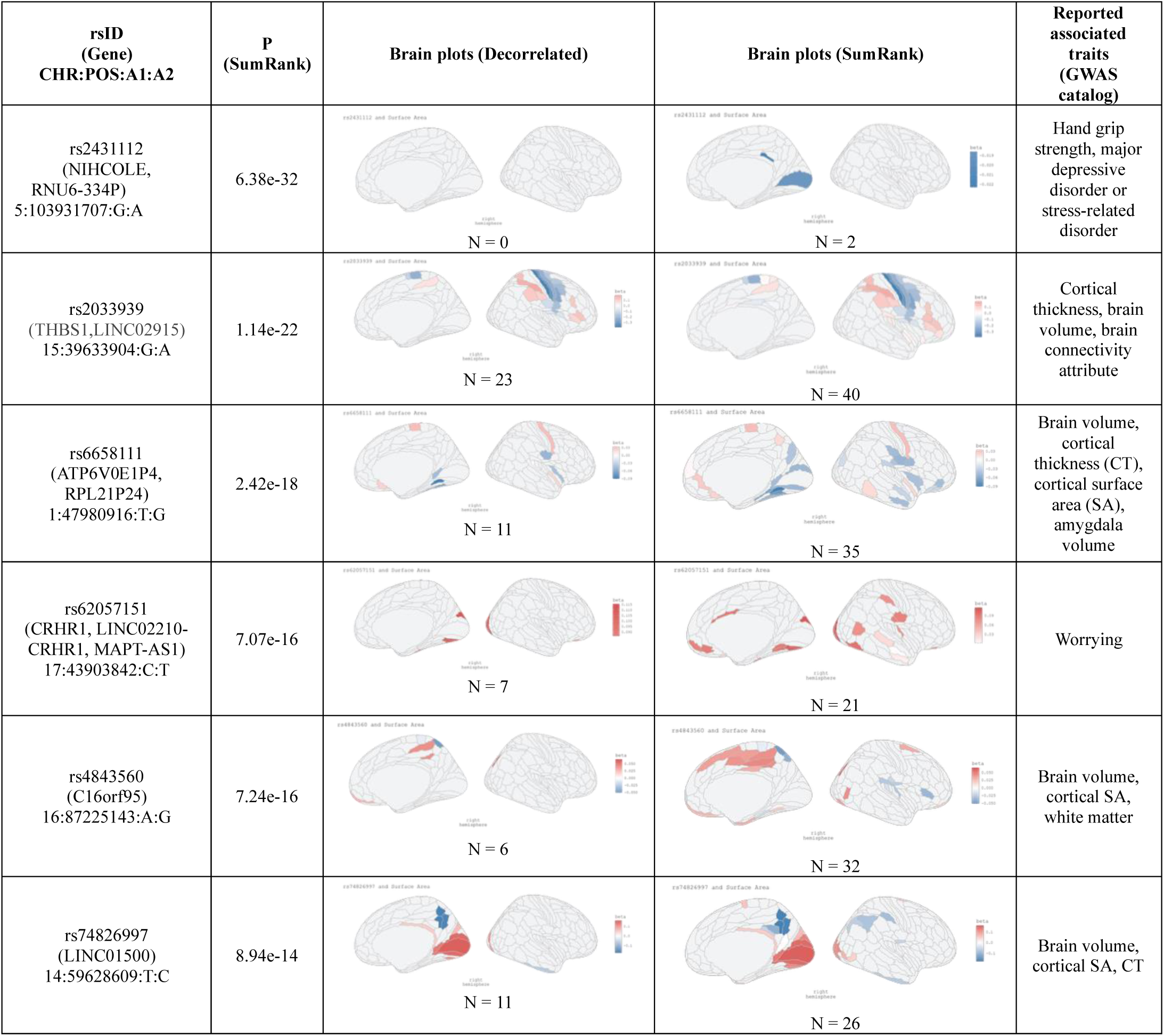
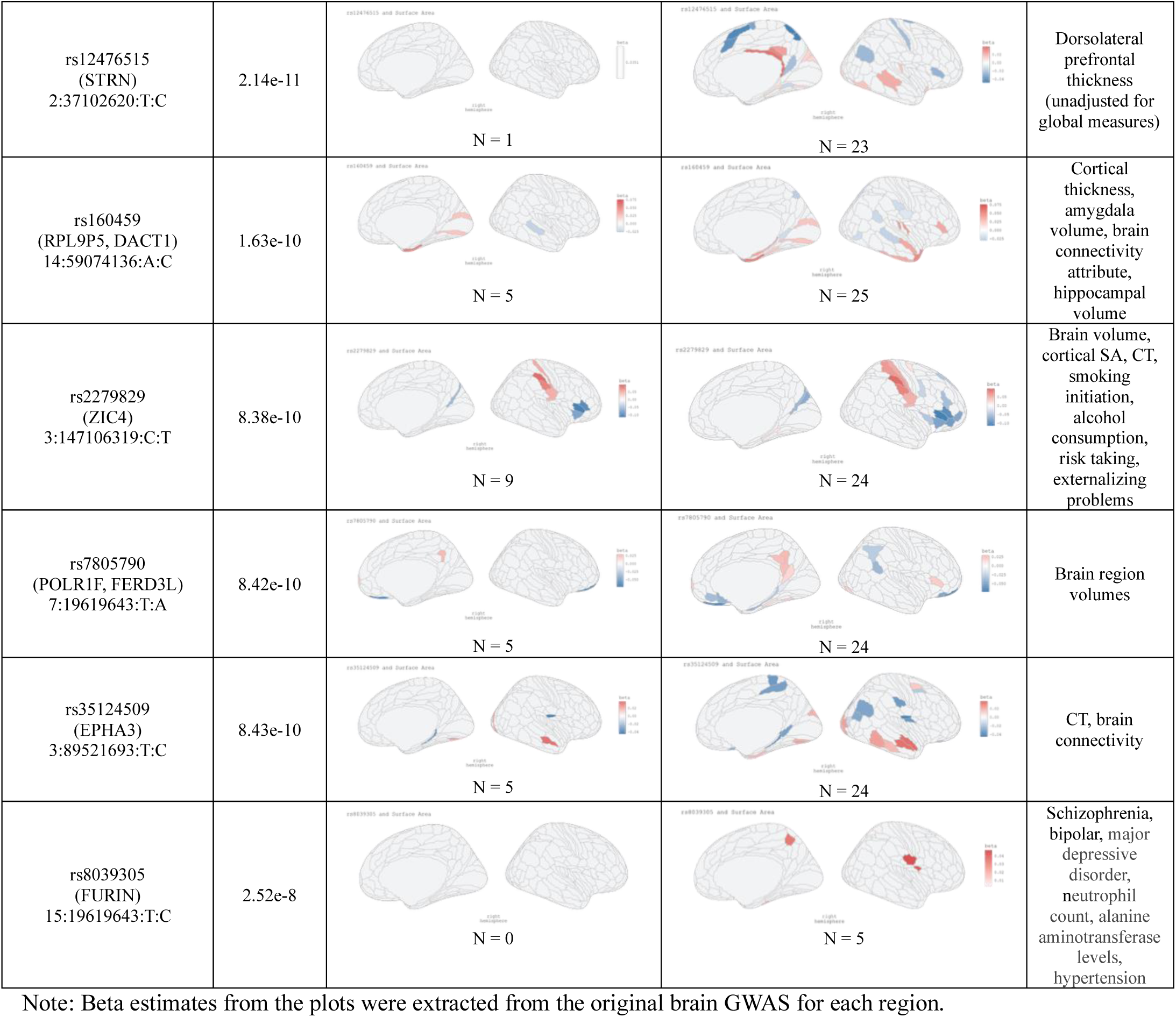
Summary of independent genomic locus (N = 12) shared with brain surface areas and all six psychiatric disorders.

| rsID<br>(Gene)<br>CHR:POS:A1:A2 | P<br>(SumRank) | Brain plots (Decorrelated) | Brain plots (SumRank) | Reported associated traits<br>(GWAS catalog) |
| --- | --- | --- | --- | --- |
| rs2431112<br>(NIHCOLE,<br>RNU6-334P)<br>5:103931707:G:A | 6.38e-32 | N = 0 | N = 2 | Hand grip strength, major depressive disorder or stress-related disorder |
| rs2033939<br>(THBS1, LINC02915)<br>15:39633904:G:A | 1.14e-22 | N = 23 | N = 40 | Cortical thickness, brain volume, brain connectivity attribute |
| rs6658111<br>(ATP6V0E1P4,<br>RPL21P24)<br>1:47980916:T:G | 2.42e-18 | N = 11 | N = 35 | Brain volume, cortical thickness (CT), cortical surface area (SA), amygdala volume |
| rs62057151<br>(CRHR1, LINC02210-CRHR1, MAPT-AS1)<br>17:43903842:C:T | 7.07e-16 | N = 7 | N = 21 | Worrying |
| rs4843560<br>(C16orf95)<br>16:87225143:A:G | 7.24e-16 | N = 6 | N = 32 | Brain volume, cortical SA, white matter |
| rs74826997<br>(LINC01500)<br>14:59628609:T:C | 8.94e-14 | N = 11 | N = 26 | Brain volume, cortical SA, CT |
| rs12476515<br>(STRN)<br>2:37102620:T:C | 2.14e-11 | rs12476515 and Surface Area<br>N = 1 | rs12476515 and Surface Area<br>N = 23 | Dorsolateral prefrontal thickness (unadjusted for global measures) |
| rs160459<br>(RPL9P5, DACT1)<br>14:59074136:A:C | 1.63e-10 | rs160459 and Surface Area<br>N = 5 | rs160459 and Surface Area<br>N = 25 | Cortical thickness, amygdala volume, brain connectivity attribute, hippocampal volume |
| rs2279829<br>(ZIC4)<br>3:147106319:C:T | 8.38e-10 | rs2279829 and Surface Area<br>N = 9 | rs2279829 and Surface Area<br>N = 24 | Brain volume, cortical SA, CT, smoking initiation, alcohol consumption, risk taking, externalizing problems |
| rs7805790<br>(POLR1F, FERD3L)<br>7:19619643:T:A | 8.42e-10 | rs7805790 and Surface Area<br>N = 5 | rs7805790 and Surface Area<br>N = 24 | Brain region volumes |
| rs35124509<br>(EPHA3)<br>3:89521693:T:C | 8.43e-10 | rs35124509 and Surface Area<br>N = 5 | rs35124509 and Surface Area<br>N = 24 | CT, brain connectivity |
| rs8039305<br>(FURIN)<br>15:19619643:T:C | 2.52e-8 | rs8039305 and Surface Area<br>N = 0 | rs8039305 and Surface Area<br>N = 5 | Schizophrenia, bipolar, major depressive disorder, neutrophil count, alanine aminotransferase levels, hypertension |

Note: Beta estimates from the plots were extracted from the original brain GWAS for each region.

Only five independent loci overlapped with general psychopathology, as defined by all six disorders, and cortical thickness of any of the 180 regions. These variants were associated with cortical thickness in 16 to 25 regions (median = 24, IQR [19,24]), with four of five variants showing divergent effects directions across cortical regions (Table 2). An exception was rs3769126 (*SLC35F6*), which demonstrated consistent negative associations with thickness across multiple cortical regions, alongside nominal positive associations with anxiety, MDD, and schizophrenia in the original GWAS analyses. These findings demonstrate the substantial directional heterogeneity of pleiotropic loci linking cortical morphology and general psychopathology and the increased discovery power of cross-trait analyses relative to single-trait analyses.

**Table 2.**
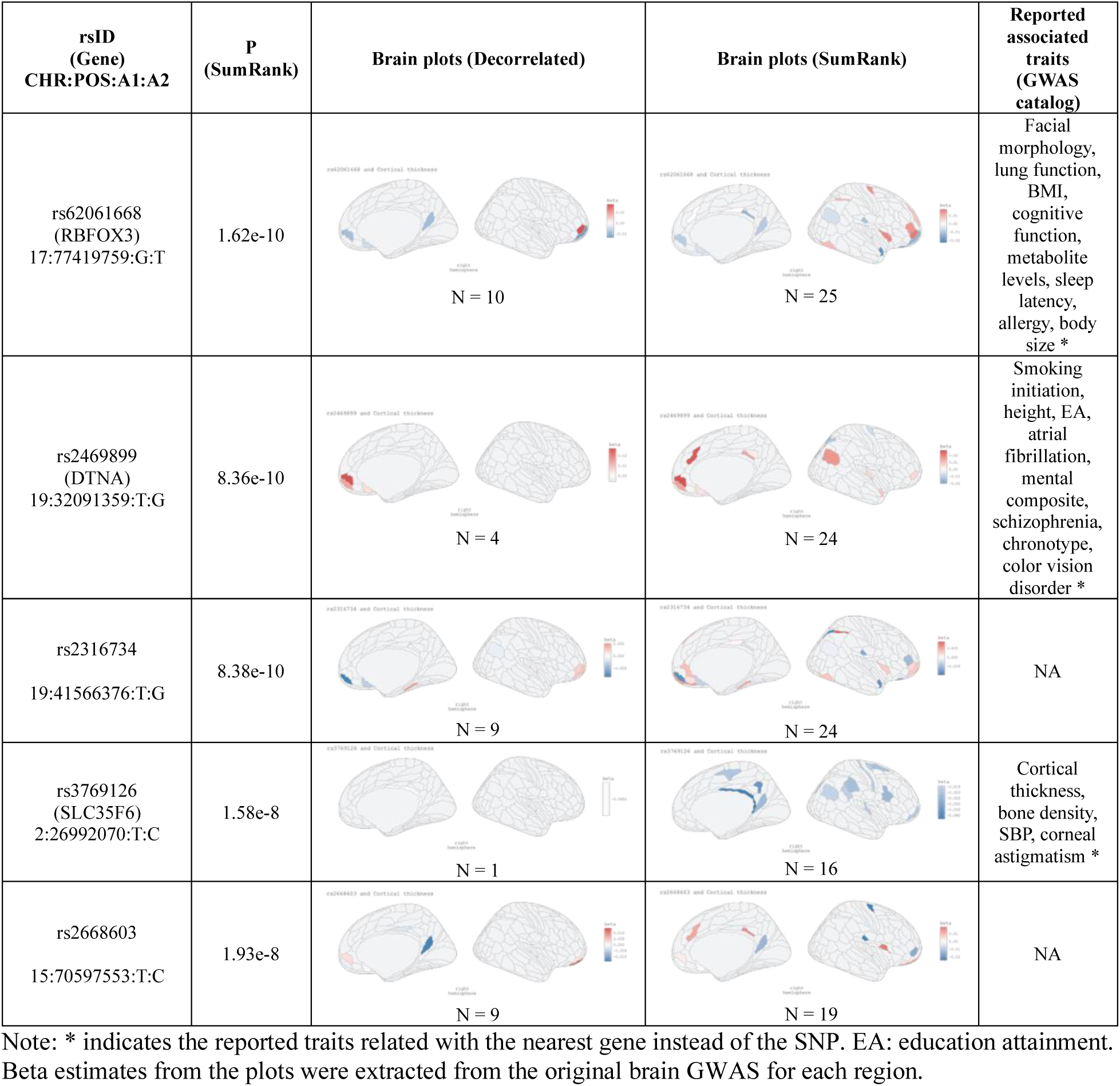
Summary of independent genomic locus (N = 5) shared with brain cortical thickness and all five psychiatric disorders.

To contextualize these findings, we assessed whether these loci had been identified in previous GWAS of brain and behavioral traits (Table 1). Of the 12 loci associated with surface area, 9 overlapped with previously reported brain measures, 3 had been associated with psychiatric or behavioral traits, and one SNP (rs2279829, mapped on *ZIC4*) had been related to both brain morphology and externalizing problems. Among the five cortical thickness loci (Table 2), three mapped to genes with established associations: rs62061668 (*RBFOX3*) has been associated with body mass index, sleep latency, and body size; rs2469899 (*DTNA*) with schizophrenia, chronotype, height, and educational attainment; and rs3769126 (*SLC35F6*) with cortical thickness, bone density, and systolic blood pressure.

### Shared biological pathways

All significant SNPs from SumRank results were further functionally annotated and mapped to genes using FUMA (v.1.7.0). To identify shared biological pathways from the shared variants, we performed gene-set analyses with the GENE2FUNC tool in FUMA. This was first done for the first 12 SumRank analyses, i.e. the six individual psychiatric disorders paired with either surface area or cortical thickness of any of the 180 brain regions. Statistically significant shared pathways were found for ADHD, anxiety, BP, MDD, and SCZ, whereas no pathways survived multiple testing correction for ASD, suggesting comparatively limited functional convergence with cortical morphology and possibly reflecting the limited GWAS discovery sample size. The 12 analyses yielded 520 statistically significant pathways, 226 being unique (Figure 3, Supplementary Data 22).

**Figure 3.**
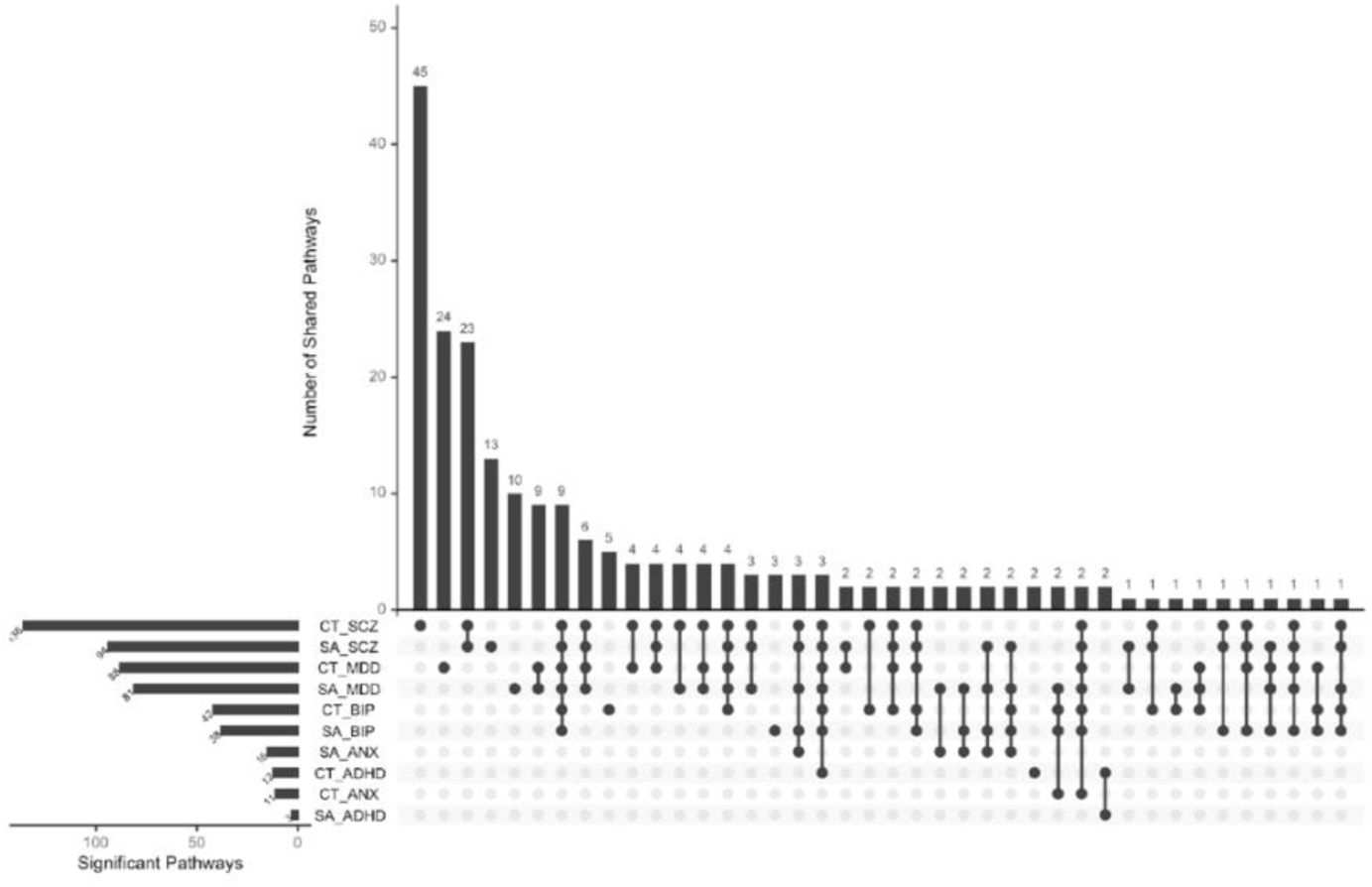
Shared Gene Ontology enrichment pathways across genetic variants shared between one or more brain structures and a specific psychiatric disorder, based on the significant findings from SumRank.

Across the 12 analyses, 9 pathways occurred most frequently, i.e. in the analyses of SCZ, MDD, and BP with both the surface area and cortical thickness of regions. These pathways converged on core neurobiological processes including synaptic signaling, regulation of trans-synaptic signaling, and neurogenesis, as well as cellular components including synapse, post-synapse, dendritic tree, axon, glutamatergic synapse, postsynaptic specialization, neuron to neuron synapse, somatodendritic compartment, and neuron projection. This highlights synaptic organization and neuronal development as linking cortical morphology with internalizing and schizophrenia/bipolar disorders. Additionally, neurogenesis and neuron development pathways were shared between cortical thickness and ADHD, while somatodendritic compartment and neuron projection were shared by cortical thickness and anxiety suggesting some disorder-specific functional overlap.

Next, we applied gene-set analyses to the results of the six cross-trait models for the three psychiatric disorder dimensions (neurodevelopmental disorders, internalizing disorders and schizophrenia/bipolar disorders) with surface area or cortical thickness. We identified 28 pathways linking brain surface area or cortical thickness with mood disorders or the combined schizophrenia/bipolar disorders group. Details were included in Supplementary Data 23). Shared enrichment included molecular function of structural constituent of chromatin, RNA binding, cellular components of DNA packaging complex, and nucleosome, implicating chromatin organization and transcriptional regulation in these pleiotropic brain and cross-disorder associations. Finally, we applied gene-set analyses to the two cross-trait associations of general psychopathology (i.e., all six disorders) together with surface area of cortical thickness. However, no significant pathways emerge.

## Discussion

We conducted the first multivariate joint analysis to identify the genetic loci shared between cortical morphology across 180 brain regions and six psychiatric disorders. Our findings revealed three key insights: (1) the overlap in causal variants between overall brain morphology and individual psychiatric disorder did not exhibit clear directional patterns except for those shared by ADHD or MDD with surface area; (2) causal variants for neurodevelopmental disorders are less likely to be involved in global cortical morphology, however, if neurodevelopmental risk loci are shared, they were associated with more widespread cortical regions than those associated with internalizing or schizophrenia/bipolar disorders; (3) most genetic loci associated with general psychopathology relate to widespread cortical regions with heterogeneous effect directions in different regions, although one locus showed highly localized effects on reduced primary visual cortex and posterior cingulate surface areas.

Our findings show a mixed directional pattern for the genetic variants related to cortical morphology and psychiatric traits. Only about half of the variants associated with the risk for a specific psychiatric disorder were related to surface area or cortical thickness in the same direction, the other half showed opposite directional influences. This pattern was only different for genetic variants influencing the risk for ADHD or MDD and total surface area; most shared causal variants displayed an opposing direction on each psychiatric disorder and total surface area. In genetic correlation estimates, a mix of concordant and discordant effects cancels out; this likely explains why genetic correlation often are low.^27,28^ Different shared genomic loci related to cortical morphology and psychiatric disorders may operate through diverse biological pathways. This may also partially explain the difficulty of identifying specific brain regions associated with psychiatric disorders. While brain morphology is moderately genetically determined, the presence of opposing effects in nearly half of the shared causal genetic variants with most psychiatric disorders complicates the characterization of the genes-brain-behavior mechanism. Further, the lack of clear directional pattern may obscure the underlying neuroanatomical biology links.

We then elucidated whether the genetic variants influencing both cortical morphology and psychiatric disorders affect the brain locally or more generally. Relatively fewer genetic risk loci for ADHD and ASD than for other disorders were related to cortical morphology. However, the few shared neurodevelopmental risk genomic loci impacted more widespread cortical regions. This finding is in line with prior findings: ADHD is characterized by widespread delayed development of cortical surface area^29^; ASD shows both altered thickness across multiple regions.^30,31^ In contrast, internalizing and schizophrenia/bipolar disorders each had a higher genetic overlap percentage with total surface area but the genomic loci were related with more localized cortical morphology. These results highlight that neurodevelopmental and mood or psychosis related disorders differ in the scale and specificity of genetic influences on cortical morphology, likely reflecting distinct neurobiological underpinnings that may contribute to clinical heterogeneity across psychiatric disorders and inform regionally targeted investigations of underlying mechanisms.

Importantly, most independent genomic loci shared by general psychopathology had widespread effects on cortical morphology, often with opposing directions across regions. This suggests heterogeneous effects of each shared locus on different cortical regions. This complex directionality pattern comes above the mixed effects of individual psychiatric disorders risk variants on overall cortical morphology. Recent research also showed that genetic variants impact widespread brain surface areas differently in specific regions.^32,33^ This reflects a complex genetic influences on brain structures that likely stems from region-specific neurodevelopmental processes and cellular diversity. It suggests that neither larger or smaller surface area and thinner or thicker cortex is a clear psychiatric risk indicator. We carefully speculate that this heterogeneity in variant effect directions across regions obscures a joint genetic signal and thus further complicates the association between overall cortical morphology and psychiatric disorders.

This is the first study identifying a genetic variant (rs2431112, near *NIHCOLE* and *RNU6-334P*) linking general psychopathology with highly specific brain regions. This variant was associated with surface area in the primary visual cortex and posterior cingulate and all six psychiatric disorders in the original GWAS and our cross-trait analyses. The primary visual cortex plays a critical role in visual working memory capacity, determining what we can hold in mind.^34^ Extensive research demonstrates that despite MDD’s heterogeneous nature, abnormal visual cortex function occurs in MDD patients and improves with antidepressant treatment.^35,36^ Similar abnormalities have been observed in schizophrenia and bipolar disorder,^37^ and primary visual cortex responses are linked to motion perception deficits in autism.^38^ The posterior cingulate cortex, a highly connected and metabolically active region, directly regulates attentional focus and has been consistently implicated in ASD, ADHD, MDD, and schizophrenia.^39^ The genetic convergence on these regions suggests potential shared pathways underlying general psychopathology. Specifically, rs2431112 may influence neurodevelopmental processes affecting both visual processing efficiency and attentional regulation, two fundamental cognitive domains disrupted across many psychiatric conditions. Both genes *NIHCOLE* and *RNU6-334P* near rs2431112 have been associated with multiple psychiatric conditions and cognition function.^40–42^

To identify the neurobiological pathways underlying the shared genetic risk loci for psychiatric disorders and brain regions, we performed functional genomic analyses. The findings converged on broad neurobiological and metabolic pathways crucial for brain development and psychiatric vulnerability, consistent with existing literature.^43,44^ Beyond transdiagnostic mechanisms, we identified disorder-specific shared pathways linking brain morphology with individual psychiatric disorders. For example, the GO cellular component of a synapse that uses GABA as a neurotransmitter is strongly supported by previous evidence for major depressive disorder (MDD)^45,46^. Additionally, odorant binding and components of the BORC complex were distinctively related to brain morphology and attention-deficit/hyperactivity disorder (ADHD). This aligns with reports linking odorant binding to autism and attention-related development, and BORC’s critical role in lysosome positioning and motility,^47,48^ with emerging evidence connecting BORC dysfunction to neurodevelopmental disorders.^47^

This study has several limitations. First, in line with common practice, the GWAS for brain regions did not adjust for global brain metrics, which may introduce confounding by overall brain size and reduce the specificity of regional associations. However, each of the 180 bilateral regions contributes only a small portion of the variance to global brain measures. Second, the ASD GWAS sample size was small and thus the limited power may have led to the null findings in the gene-set analyses. Third, the SumRank method incorporates only p-values and lacks effect size information, which may have limited the insight into the biological and functional impact of some variants, especially in the presence of heterogeneous effect sizes. Nonetheless, most cross-trait methods similarly rely on p-value based input, and p-values are simpler to interpret and compare across traits. Fourth, GWAS for psychiatric disorders may include participants with unaccounted comorbidities and thus capture more pleiotropy, and case heterogeneity may introduce noise into the genetic signals.

Taken together, we identified numerous independent genomic loci shared between brain morphology and individual psychiatric disorders, as well as many genomic loci shared across psychiatric cross-disorder dimensions or general psychopathology. We found a notable locus linked to multiple psychiatric disorders, specifically associated with the primary visual and posterior cingulate cortices. In general, however, the genetic architecture linking cortical morphology with psychiatric disorders is characterized by pronounced heterogeneity in the direction and anatomical distribution of these shared effects. Rather than exerting uniform influences, most shared variants displayed opposing associations across brain regions, with individual SNPs increasing surface area or thickness in some regions while decreasing it in others. This highly non-uniform pattern observed across most individual disorders except ADHD and MDD, across diagnostic dimensions, and across loci shared with general psychopathology suggests that risk variants act through multiple, region-specific neurodevelopmental and neurobiological pathways. Consequently, broader measures of cortical morphology are unlikely to serve as endophenotypes for psychiatric disorders, as nearly half the loci shared by most psychiatric disorders and cortical structures showed opposing effects. Moreover, the pleiotropic locus shared by all psychiatric disorders predominately each showed opposing effects across different cortical regions. These effects likely obscure some meaningful relationships at the whole-brain level.

## Online Methods

### Summary Statistics

We used GWAS summary statistics^49^ of surface area and cortical thickness of 180 bilaterally averaged brain regions, defined by the Human Connectome Project Multi-Modal Parcellation version 1.0.^50^ This map parcellation was based on multi-modal MRI data, including structural, functional, and connectivity features. These GWAS of 180 brain regions used data from both the UK Biobank (N = 31,797) and ABCD (N = 4,866) cohorts. The genetic information of the psychiatric disorders was derived from the most recent GWAS summary statistics from the Psychiatric Genomics Consortium: anxiety (N cases = 96,888, N controls = 671,768),^8^ ADHD (N cases = 38,691, N controls= 186,843),^51^ ASD (N cases = 18,381, N controls = 27,969),^52^ BP (N cases = 57,833, N controls = 722,909),^53^ MDD (N cases = 357,636, N controls = 1,281,936),^54^ and schizophrenia (N cases = 53,386, N controls = 77,258).^55^ To minimize sample overlap and reduce inflation from cross-trait enrichment, UK Biobank participants were excluded from the psychiatric GWAS. All individuals included in the analyses were of European ancestry because of the lack of non-European GWAS of regional brain structures. We included overlapping genetic variants across brain region GWAS and psychiatric disorders and thus 5,762,210 and 5,787,282 variants remained in the analyses for surface area and cortical thickness separately. Given that the GWAS summary statistics for the 180 brain regions were derived from the same sample, we applied a whitening transformation via Cholesky decomposition of the z-score covariance matrix to decorrelate the z-statistics and mitigate bias due to sample overlap.

### Genetic overlap between overall brain structures and each psychiatric disorder

To characterize the shared genetic architecture between overall brain structure measures, total surface area and cortical thickness, and psychiatric disorders, we applied causal mixture models using MiXeR (v1.3) (https://github.com/precimed/mixer). Univariate MiXeR analyses estimated each trait’s polygenicity (i.e. the number of causal variants explaining 90% of SNP heritability) and discoverability (i.e. the average effect size of causal variants).^56^ Bivariate MiXeR models were then used to quantify the number of shared and trait-specific causal variants for each trait pair, as well as the proportion of shared variants with concordant effect directions.^28^ We quantified the proportion of SNPs of the overall brain structures overlapping with each psychiatric disorders and calculated the genetic correlation within the shared genetic components. In line with recommendations, the models were run on 20 random subsets of ∼600K SNPs from the 1000 Genomes EUR panel (phase 3 version 5) excluding the MHC region (chromosome 6 position between 25726291 and 33377699 in the hg19 reference), and the model parameters were averaged across the 20 runs.

### Cross-trait associations

Next, we aimed to identify genetic variants shared between psychiatric disorders and cortical regions. We used SumRank, a method that integrates GWAS p-values across multiple traits to determine, for each variant, the subset of traits to which it is most strongly associated.^25^ Sumrank computes a new p-value for every possible subset of traits and subsequently selecting the lowest p-value. Previous work shows that SumRank robustly controls the false positive rate while being well-powered to detect cross-trait associations across hundreds of traits. We applied SumRank to the summary statistics for six psychiatric traits and the cortical thickness and surface area of 180 brain regions. The analyses were conducted in three stages.

First, we conducted disorder-specific analyses, using SumRank to identify genetic variants associated with each psychiatric disorder and at least one cortical region, separately for thickness and surface area. Second, to investigate broader trans-diagnostic patterns, we grouped the psychiatric disorders according to previously established genomic dimensions from Genomic Structural Equation Modeling: neurodevelopmental disorders (ASD and ADHD), internalizing disorders (anxiety and MDD), and schizophrenia/bipolar disorders (SCZ/BP).^57^ We reapplied SumRank separately for each group, to identify the shared genetic variants associated with the disorder group and at least one cortical region. Third, we assessed the shared genetic association of general psychopathology (i.e. all 6 disorders) with regional or global cortical structures by constraining SumRank to examine only subsets which included all six psychiatric disorders and at least one brain region. SNPs with SumRank p-values < 5 × 10⁻⁸ were considered statistically significant pleiotropic variants. To control the false positive rate, we filtered out p-values above the threshold 3.9e-4, calculate by 1/(*m*√*m*), where *m* is the number of traits included (m=186). This threshold tended to work best in avoiding false positives in the simulations.

### Independent genomic risk loci

Independent significant SNPs were defined using an LD block merging distance ≤250 kb, r² < 0.6, based on the 1000 Genomes EUR panel. Among these, we defined lead SNPs as those with r² < 0.1, with each locus represented by the lead SNP with the lowest SumRank p-value. To characterize the distribution of brain regions associated with each independent genomic locus and general psychopathology, we extracted effect size estimates for lead SNPs from their original GWASs under two p-value thresholds. First, we obtained estimates for brain regions reaching genome-wide significance in the decorrelated matrix from original GWAS (p < 5×10⁻⁸). Second, we extracted estimates for brain regions based on SumRank cross-trait p-values (p < 5×10⁻⁸). This allowed us to examine brain region patterns linked to general psychopathology for each shared genomic locus and to identify novel associations from cross-trait analyses that were not detected in the original GWAS. To contextualize our findings and compare with existing findings, we also linked each independent genomic loci with previously reported associated traits based on the GWAS catalog. For the lead SNPs that have not been found to be associated with any traits, we reported the traits linked with their mapped genes.

### Functional annotation

All significant SNPs from SumRank results were further functionally annotated using FUMA (v.1.7.0), including functional consequences (Ensemble v102 with ANNOVAR). Gene mapping was also performed using FUMA by employing three approaches: positional mapping, expression quantitative locus (eQTL) mapping, and chromatin interaction mapping. For positional mapping, variants were mapped to any protein-coding gene that was within 10 kb. For eQTL mapping, variants were mapped within a 1 Mb region to the cis-eQTL expression of genes based on the BRAINEAC 11 tissues including cerebellar cortex, frontal cortex, hippocampus, inferior olivary nucleus, occipital cortex, putamen, substantial nigra, temporal cortex, thalamus, intralobular white matter and averaged expression of 10 brain regions. The Benjamini-Hochberg procedure was used to adjust the false discovery rate of the eQTL mapping to 0.05. For chromatin interaction mapping, variants were linked to transcription start sites of genes based on chromatin loops that enable DNA-DNA interactions in 3D space. Data from HiC (Giusti-Rodriguez et al., 2019) was used for both adult and fetal cortex tissues. Chromatin interaction mapping was further constrained to genetic variants in enhancers or promotors based on the 11 brain-related roadmaps from the NIH Roadmap Epigenomics Consortium project. Again, the Benjamini-Hochberg procedure was used to adjust p-values to control the false discovery rate to1E-6, in line with recommendations.

Using the GENE2FUNC module in FUMA, we performed gene-set analyses across 12 combinations, each pairing one of six individual psychiatric disorders with one or more brain surface area or cortical thickness regions. We also conducted additional analyses: six combinations involving the three psychiatric disorder dimensions, and two combinations including all six disorders together with the brain regions. Hypergeometric tests were used to test whether Gene Ontology (GO) gene sets were enriched for the mapped genes. The Benjamini-Hochberg procedure was used to adjust the p-values and set the false discovery rate to 0.05.

## Data Availability

All data produced in the present study are available upon reasonable request to the authors.

## Acknowledgements

T.G. is supported by NIMH R01MH130899.

## Contributions

Y.Z., H.T. and S.L. jointly conceived of the project and designed the analyses. Y.Z. and S.L. carried out analysis and visualization. Anxiety Disorders Working Group of the Psychiatric Genomics Consortium contributed data. Y.Z., T.G., T.T.T., K.W.C., H.T. and S.L. contributed to interpretation. H.T. and S.L. supervised the project. Y.Z. wrote the paper, with feedback from all co-authors. All authors approved the paper.

## References

1. Kendell R, Jablensky A. Distinguishing Between the Validity and Utility of Psychiatric Diagnoses. American Journal of Psychiatry. 2003/01/01 2003;160(1):4–12. doi:10.1176/appi.ajp.160.1.4

2. Smoller JW. Disorders and borders: Psychiatric genetics and nosology. American Journal of Medical Genetics Part B: Neuropsychiatric Genetics. 2013/10/01 2013;162(7):559–578. 10.1002/ajmg.b.32174

3. Borsboom D, Cramer AO, Schmittmann VD, Epskamp S, Waldorp LJ. The small world of psychopathology. PLoS One. 2011;6(11):e27407. doi:10.1371/journal.pone.0027407

4. Kessler RC, Chiu WT, Demler O, Merikangas KR, Walters EE. Prevalence, severity, and comorbidity of 12-month DSM-IV disorders in the National Comorbidity Survey Replication. Arch Gen Psychiatry. Jun 2005;62(6):617–27. doi:10.1001/archpsyc.62.6.617

5. Grotzinger AD, Werme J, Peyrot WJ, et al. Mapping the genetic landscape across 14 psychiatric disorders. Nature. 2026/01/01 2026;649(8096):406–415. doi:10.1038/s41586-025-09820-3

6. Andreassen OA, Hindley GFL, Frei O, Smeland OB. New insights from the last decade of research in psychiatric genetics: discoveries, challenges and clinical implications. World Psychiatry. Feb 2023;22(1):4–24. doi:10.1002/wps.21034

7. Friligkou E, Løkhammer S, Cabrera-Mendoza B, et al. Gene discovery and biological insights into anxiety disorders from a large-scale multi-ancestry genome-wide association study. Nature Genetics. 2024/10/01 2024;56(10):2036–2045. doi:10.1038/s41588-024-01908-2

8. Strom NI, Verhulst B, Bacanu S-A, et al. Genome-wide association study of major anxiety disorders in 122,341 European-ancestry cases identifies 58 loci and highlights GABAergic signaling. Nature Genetics. 2026/02/03 2026;doi:10.1038/s41588-025-02485-8

9. Genomic Relationships, Novel Loci, and Pleiotropic Mechanisms across Eight Psychiatric Disorders. Cell. Dec 12 2019;179(7):1469–1482.e11. doi:10.1016/j.cell.2019.11.020

10. Bourque VR, Poulain C, Proulx C, et al. Genetic and phenotypic similarity across major psychiatric disorders: a systematic review and quantitative assessment. Transl Psychiatry. Mar 30 2024;14(1):171. doi:10.1038/s41398-024-02866-3

11. Grotzinger AD. Shared genetic architecture across psychiatric disorders. Psychol Med. Oct 2021;51(13):2210–2216. doi:10.1017/s0033291721000829

12. Lee PH, Feng YA, Smoller JW. Pleiotropy and Cross-Disorder Genetics Among Psychiatric Disorders. Biol Psychiatry. Jan 1 2021;89(1):20–31. doi:10.1016/j.biopsych.2020.09.026

13. Bourque V-R, Poulain C, Proulx C, et al. Genetic and phenotypic similarity across major psychiatric disorders: a systematic review and quantitative assessment. Translational Psychiatry. 2024/03/30 2024;14(1):171. doi:10.1038/s41398-024-02866-3

14. Williams CM, Peyre H, Wolfram T, et al. Characterizing the phenotypic and genetic structure of psychopathology in UK Biobank. Nature Mental Health. 2024/08/01 2024;2(8):960–974. doi:10.1038/s44220-024-00272-8

15. Marek S, Tervo-Clemmens B, Calabro FJ, et al. Reproducible brain-wide association studies require thousands of individuals. Nature. 2022/03/01 2022;603(7902):654–660. doi:10.1038/s41586-022-04492-9

16. Disorder WCftA-DH, Disorder AS, Disorder B, Disorder MD, Disorder O-C, Groups aSEW. Virtual Histology of Cortical Thickness and Shared Neurobiology in 6 Psychiatric Disorders. JAMA Psychiatry. 2021;78(1):47–63. doi:10.1001/jamapsychiatry.2020.2694

17. Opel N, Goltermann J, Hermesdorf M, Berger K, Baune BT, Dannlowski U. Cross-Disorder Analysis of Brain Structural Abnormalities in Six Major Psychiatric Disorders: A Secondary Analysis of Mega– and Meta-analytical Findings From the ENIGMA Consortium. Biol Psychiatry. Nov 1 2020;88(9):678–686. doi:10.1016/j.biopsych.2020.04.027

18. Matsumoto J, Fukunaga M, Miura K, et al. Cerebral cortical structural alteration patterns across four major psychiatric disorders in 5549 individuals. Molecular Psychiatry. 2023/11/01 2023;28(11):4915–4923. doi:10.1038/s41380-023-02224-7

19. Scarpazza C, Ha M, Baecker L, et al. Translating research findings into clinical practice: a systematic and critical review of neuroimaging-based clinical tools for brain disorders. Transl Psychiatry. Apr 20 2020;10(1):107. doi:10.1038/s41398-020-0798-6

20. Li Z, Li D, He Y, Wang K, Ma X, Chen X. Cross-Disorder Analysis of Shared Genetic Components Between Cortical Structures and Major Psychiatric Disorders. Schizophr Bull. Sep 1 2022;48(5):1145–1154. doi:10.1093/schbul/sbac019

21. Radonjić NV, Hess JL, Rovira P, et al. Structural brain imaging studies offer clues about the effects of the shared genetic etiology among neuropsychiatric disorders. Mol Psychiatry. Jun 2021;26(6):2101–2110. doi:10.1038/s41380-020-01002-z

22. Stauffer E-M, Bethlehem RAI, Dorfschmidt L, Won H, Warrier V, Bullmore ET. The genetic relationships between brain structure and schizophrenia. Nature Communications. 2023/11/28 2023;14(1):7820. doi:10.1038/s41467-023-43567-7

23. Cheng W, van der Meer D, Parker N, et al. Shared genetic architecture between schizophrenia and subcortical brain volumes implicates early neurodevelopmental processes and brain development in childhood. Mol Psychiatry. Dec 2022;27(12):5167–5176. doi:10.1038/s41380-022-01751-z

24. Sha Z, Warrier V, Bethlehem RAI, et al. The overlapping genetic architecture of psychiatric disorders and cortical brain structure. Nature Mental Health. 2025/09/01 2025;3(9):1020–1036. doi:10.1038/s44220-025-00475-7

25. Lamballais S, Roshchupkin GV, Poot RA, et al. A comprehensive benchmarking and validation study of cross-trait association methods. medRxiv. 2025:2022.09.07.22279671. doi:10.1101/2022.09.07.22279671

26. Nagel M, Watanabe K, Stringer S, Posthuma D, van der Sluis S. Item-level analyses reveal genetic heterogeneity in neuroticism. Nature Communications. 2018/03/02 2018;9(1):905. doi:10.1038/s41467-018-03242-8

27. Hindley G, Frei O, Shadrin AA, et al. Charting the Landscape of Genetic Overlap Between Mental Disorders and Related Traits Beyond Genetic Correlation. Am J Psychiatry. Nov 1 2022;179(11):833–843. doi:10.1176/appi.ajp.21101051

28. Frei O, Holland D, Smeland OB, et al. Bivariate causal mixture model quantifies polygenic overlap between complex traits beyond genetic correlation. Nature Communications. 2019/06/03 2019;10(1):2417. doi:10.1038/s41467-019-10310-0

29. Yadav SK, Bhat AA, Hashem S, et al. Genetic variations influence brain changes in patients with attention-deficit hyperactivity disorder. Translational Psychiatry. 2021/06/05 2021;11(1):349. doi:10.1038/s41398-021-01473-w

30. Ecker C, Bookheimer SY, Murphy DGM. Neuroimaging in autism spectrum disorder: brain structure and function across the lifespan. The Lancet Neurology. 2015;14(11):1121–1134. doi:10.1016/S1474-4422(15)00050-2

31. Hashem S, Nisar S, Bhat AA, et al. Genetics of structural and functional brain changes in autism spectrum disorder. Translational Psychiatry. 2020/07/13 2020;10(1):229. doi:10.1038/s41398-020-00921-3

32. Segal A, Parkes L, Aquino K, et al. Regional, circuit and network heterogeneity of brain abnormalities in psychiatric disorders. Nature Neuroscience. 2023/09/01 2023;26(9):1613–1629. doi:10.1038/s41593-023-01404-6

33. Sha Z, Warrier V, Bethlehem RAI, et al. The overlapping genetic architecture of psychiatric disorders and cortical brain structure. Nature Mental Health. 2025/08/11 2025;doi:10.1038/s44220-025-00475-7

34. Espinosa JS, Stryker MP. Development and plasticity of the primary visual cortex. Neuron. 2012;75(2):230–249.

35. Wu F, Lu Q, Kong Y, Zhang Z. A Comprehensive Overview of the Role of Visual Cortex Malfunction in Depressive Disorders: Opportunities and Challenges. Neuroscience Bulletin. 2023/09/01 2023;39(9):1426–1438. doi:10.1007/s12264-023-01052-7

36. Zhou R, Wang F, Zhao G, et al. Effects of tumor necrosis factor-α polymorphism on the brain structural changes of the patients with major depressive disorder. Translational Psychiatry. 2018/10/11 2018;8(1):217. doi:10.1038/s41398-018-0256-x

37. Reavis EA, Lee J, Wynn JK, Engel SA, Jimenez AM, Green MF. Cortical thickness of functionally defined visual areas in schizophrenia and bipolar disorder. Cerebral Cortex. 2017;27(5):2984–2993.

38. Robertson CE, Thomas C, Kravitz DJ, et al. Global motion perception deficits in autism are reflected as early as primary visual cortex. Brain. 2014;137(9):2588–2599. doi:10.1093/brain/awu189

39. Leech R, Sharp DJ. The role of the posterior cingulate cortex in cognition and disease. Brain. 2014;137(1):12–32. doi:10.1093/brain/awt162

40. Lee JJ, Wedow R, Okbay A, et al. Gene discovery and polygenic prediction from a genome-wide association study of educational attainment in 1.1 million individuals. Nature Genetics. 2018/08/01 2018;50(8):1112–1121. doi:10.1038/s41588-018-0147-3

41. Peyrot WJ, Price AL. Identifying loci with different allele frequencies among cases of eight psychiatric disorders using CC-GWAS. Nature Genetics. 2021/04/01 2021;53(4):445–454. doi:10.1038/s41588-021-00787-1

42. Wu Y, Cao H, Baranova A, et al. Multi-trait analysis for genome-wide association study of five psychiatric disorders. Translational Psychiatry. 2020/07/19 2020;10(1):209. doi:10.1038/s41398-020-00902-6

43. Khan Y, Davis CN, Jinwala Z, et al. Transdiagnostic and Disorder-Level Genome-Wide Association Studies Enhance Precision of Substance Use and Psychiatric Genetic Risk Profiles in African and European Ancestries. Biological Psychiatry. 2025/05/08/ 2025; 10.1016/j.biopsych.2025.04.021

44. Taylor JJ, Lin C, Talmasov D, et al. A transdiagnostic network for psychiatric illness derived from atrophy and lesions. Nat Hum Behav. Mar 2023;7(3):420–429. doi:10.1038/s41562-022-01501-9

45. Fogaça MV, Duman RS. Cortical GABAergic Dysfunction in Stress and Depression: New Insights for Therapeutic Interventions. Review. Frontiers in Cellular Neuroscience. 2019–March–12 2019; Volume 13 – 2019 doi:10.3389/fncel.2019.00087

46. Luscher B, Shen Q, Sahir N. The GABAergic deficit hypothesis of major depressive disorder. Molecular psychiatry. 2011;16(4):383–406.

47. Hartwig C, Monis WJ, Chen X, Dickman DK, Pazour GJ, Faundez V. Neurodevelopmental disease mechanisms, primary cilia, and endosomes converge on the BLOC-1 and BORC complexes. Developmental Neurobiology. 2018/03/01 2018;78(3):311–330. 10.1002/dneu.22542

48. Wang J, Zhang Q, Fan W, et al. Deciphering olfactory receptor binding mechanisms: a structural and dynamic perspective on olfactory receptors. Review. Frontiers in Molecular Biosciences. 2025–January–08 2025;Volume 11 – 2024doi:10.3389/fmolb.2024.1498796

49. Warrier V, Zhang X, Reed P, et al. Genetic correlates of phenotypic heterogeneity in autism. Nature Genetics. 2022/09/01 2022;54(9):1293–1304. doi:10.1038/s41588-022-01072-5

50. Glasser MF, Coalson TS, Robinson EC, et al. A multi-modal parcellation of human cerebral cortex. Nature. 2016/08/01 2016;536(7615):171–178. doi:10.1038/nature18933

51. Demontis D, Walters GB, Athanasiadis G, et al. Genome-wide analyses of ADHD identify 27 risk loci, refine the genetic architecture and implicate several cognitive domains. Nat Genet. Feb 2023;55(2):198–208. doi:10.1038/s41588-022-01285-8

52. Grove J, Ripke S, Als TD, et al. Identification of common genetic risk variants for autism spectrum disorder. Nat Genet. Mar 2019;51(3):431–444. doi:10.1038/s41588-019-0344-8

53. O’Connell KS, Koromina M, van der Veen T, et al. Genomics yields biological and phenotypic insights into bipolar disorder. Nature. Mar 2025;639(8056):968–975. doi:10.1038/s41586-024-08468-9

54. Trans-ancestry genome-wide study of depression identifies 697 associations implicating cell types and pharmacotherapies. Cell. Feb 6 2025;188(3):640–652.e9. doi:10.1016/j.cell.2024.12.002

55. Trubetskoy V, Pardiñas AF, Qi T, et al. Mapping genomic loci implicates genes and synaptic biology in schizophrenia. Nature. Apr 2022;604(7906):502–508. doi:10.1038/s41586-022-04434-5

56. Holland D, Frei O, Desikan R, et al. Beyond SNP heritability: Polygenicity and discoverability of phenotypes estimated with a univariate Gaussian mixture model. PLOS Genetics. 2020;16(5):e1008612. doi:10.1371/journal.pgen.1008612

57. Grotzinger AD, Mallard TT, Akingbuwa WA, et al. Genetic architecture of 11 major psychiatric disorders at biobehavioral, functional genomic and molecular genetic levels of analysis. Nat Genet. May 2022;54(5):548–559. doi:10.1038/s41588-022-01057-4

