## Supplementary Figure 1 for "Shared genetic architecture of cortical morphology and psychiatric disorders: insights from a cross-trait analyses across 180 cortical regions"

Supplemental Figure 1 Distribution of genetic variants shared by a certain number of brain regions (cortical thickness and area) with psychiatric traits: Different patterns for specific psychiatric disorders, psychiatric disorder groups, and general psychopathology.

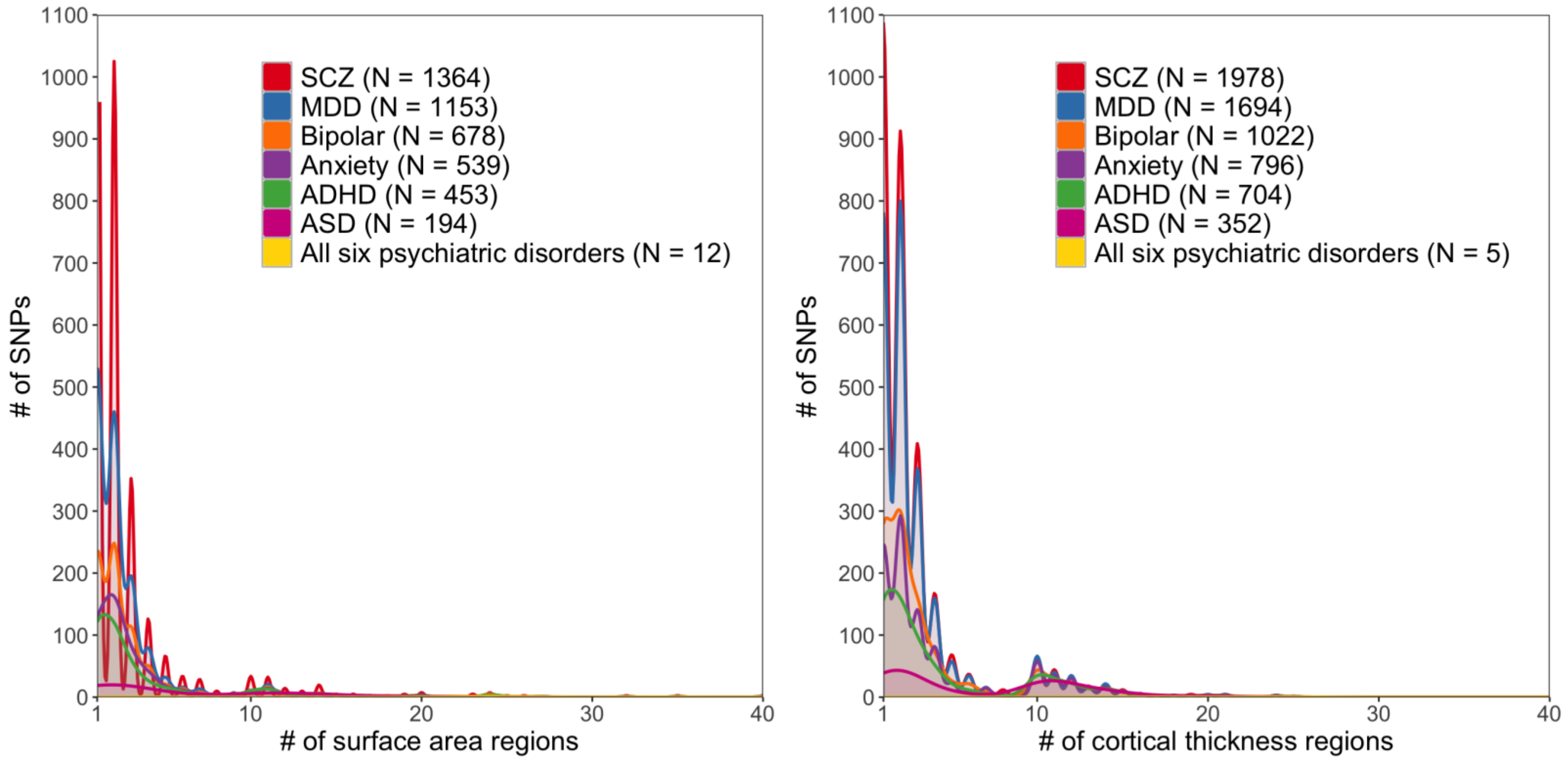

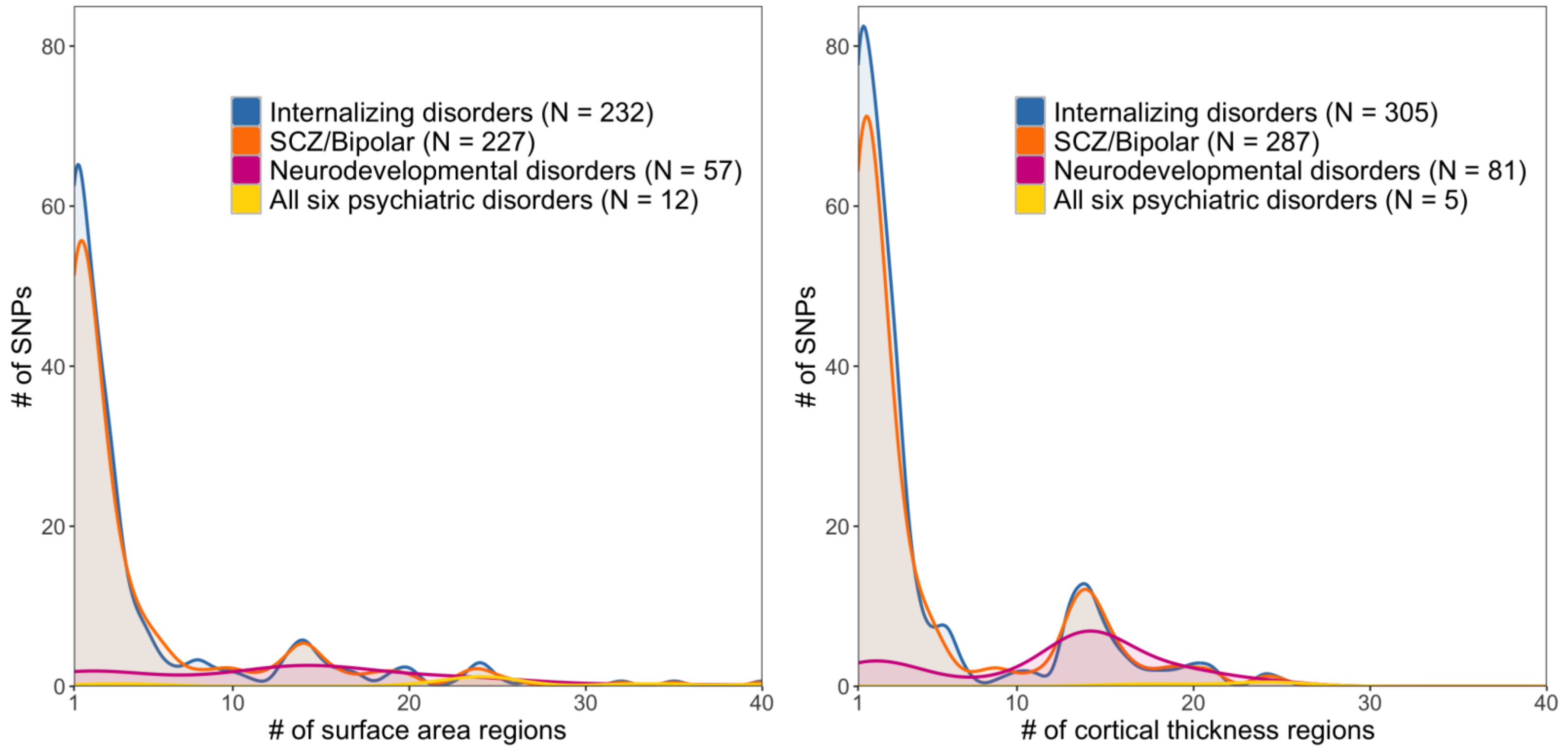

Internalizing disorders (N = 232)
SCZ/Bipolar (N = 227)
Neurodevelopmental disorders (N = 57)
All six psychiatric disorders (N = 12)
### of SNPs
### of surface area regions
Internalizing disorders (N = 305)
SCZ/Bipolar (N = 287)
Neurodevelopmental disorders (N = 81)
All six psychiatric disorders (N = 5)
### of SNPs
### of cortical thickness regions
